# Analysis and Prediction of COVID-19 Characteristics Using a Birth-and-Death Model

**DOI:** 10.1101/2020.06.23.20138719

**Authors:** Narayanan C. Viswanath

## Abstract

Its spreading speed together with the risk of fatality might be the main characteristic that separates COVID-19 from other infectious diseases in our recent history. In this scenario, mathematical modeling for predicting the spread of the disease could have great value in containing the disease. Several very recent papers have contributed to this purpose. In this study we propose a birth-and-death model for predicting the number of COVID-19 active cases. It relation to the Susceptible-Infected-Recovered (SIR) model has been discussed. An explicit expression for the expected number of active cases helps us to identify a stationary point on the infection curve, where the infection ceases increasing. Parameters of the model are estimated by fitting the expressions for active and total reported cases simultaneously. We analyzed the movement of the stationary point and the basic reproduction number during the infection period up to the 20^th^ of April 2020. These provide information about the disease progression path and therefore could be really useful in designing containment strategies.

## Introduction

Mathematical modeling of the spread of an infectious disease has a vast history dating back to 1760 when Daniel Bernoulli first modeled the spread of smallpox [1]. Among popular models, there is the Susceptible-Infected-Recovered (SIR) model first proposed by A. G. McKendrick and W. O. Kermack in 1927 [2]. Since then, several authors have studied SIR models and its variants. A detailed account of such studies can be found in [3]. Since these models include three variables with nonlinear relations between them, analytical solution has not been obtained for the general SIR model; though it is done for some special cases [3, 4]. The Susceptible-Exposed-Infected-Recovered (SEIR) model is an extension of the SIR model. Growth models such as logistic [5] and Gompertz [6] have the ability to capture the point where growth stops or become very slow. These are therefore successful in modeling many long term real world data including that of epidemic.

Prem et al. [7], Fang et al. [8], Kuniya [9] and Lin et al. [10] are among the very recent papers which applied the SEIR model for modeling the COVID-19 disease progression and its future path prediction. [7] studies the effect of physical distancing measures in reducing COVID-19 infection peak. [8] performs a fitting of the active cases in China and observed that its peak arrived on 15 February 2020. [9] studies the epidemic size and its relation to the physical interventions in Japan. The effect of individual reaction and governmental action is studied in [10]. Anastassopoulou et al. [11] applies a Susceptible-Infected-Recovered/Dead (SIRD) model for forecasting the COVID-19 outbreak. They obtain the basic reproduction number *R*_0_. Using a stochastic transmission model, Kucharski et al. [12] computes *R*_0_. Zhang et al. [13] computes *R*_0_ on the basis of a branching process model.

Unavailability of explicit expression for quantities such as actively infected cases may be considered as a drawback of the SIR and its extension models. In this paper, we propose a simplified model for predicting the number of persons actively infected during the spread of COVID-19. We consider a birth-and-death model with time dependent birth rate [14] for this purpose. It turns out that this is a simplified version of the SIR model. We hope to estimate the parameters of the model by fitting it to the real data and hence to justify that the simplification has indeed served its purpose.

## Methods

Let *AI*(*t*) denote the number of COVID-19 infected active cases at time *t*. Then {*AI*(*t*)} may be regarded as a birth and death process [8] with birth rate *λ* (*t*) (production rate of infections) and recovery rate µ. Here the recovery rate µ represents the rate at which an infected person either dies or gets cured. Let *I*(*t*) be the average value of *AI*(*t*) and *R*(*t*) be the average value of recovered/dead cases up to time *t*. Let *M*(*t*) be the sum of *I*(*t*) and *R*(*t*). Then *M*(*t*) is the total number of cases reported up to time *t*. It is known that *I*(*t*) and *M*(*t*) satisfies the differential equations (please refer [14]):

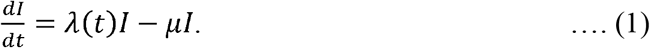

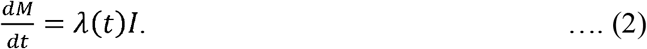

It then follows from equations (1) and (2) that

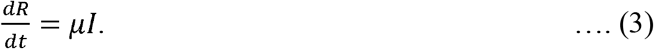

Now consider an SIR model given by the set of differential equations

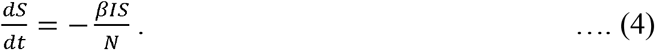

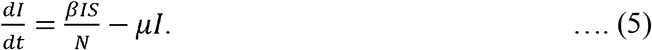

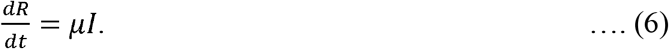

On comparison we find that equation (1) can be obtained from equation (5) by assuming *λ* (*t*) as equal to 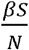, where *N* is the population size. Since *s*(*t*) is equal to *N*− (*I*(*t*) + *R*(*t*)), we can find the number of susceptible cases from equations (1-3) as *N*− *M*(*t*). The basic reproduction number is given by:

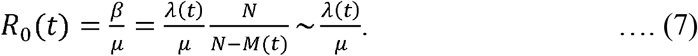

Notice that we have taken the basic reproduction number *R*_0_ as a function of time; whereas in a standard SIR model it is a constant. Naturally, the overall infection production rate has to decrease with time for the disease to suppress. In the standard SIR model, this is attained by assuming that the overall disease production rate is 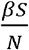 (please refer equation (5)). However *s*(*t*), the number of susceptible people, is a variable about which very less real information is available. Hence it may not be possible to apply a fitting of the real data to this variable. Considering this fact, our choice of the *R*_0_ as equal to 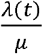 may be justified if we were able to fit this to real data.

Now we assume that *λ* (*t*) = *ae*^−*bt*^, to reflect the fact that the infection production rate may decrease with time as more people goes on isolation as time progresses. It then follows from [14] that equations (1-3) can be solved to give:

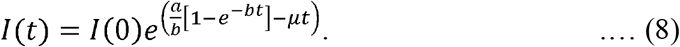

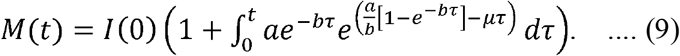

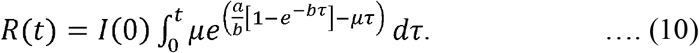

It follows from (8) that the stationary point on the infection curve, which is the graph of *I*(*t*), occurs at the time point

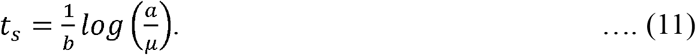

*t*_*s*_ can be thought of as the point where the active infection count stops increasing. The time varying basic reproduction number *R*_0_(*t*) is given by:

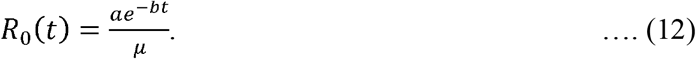

In the case of COVID-19, we have the data of active cases and total cases. The parameters *I*(0), *a, h* and *µ* were estimated by fitting the curves given by equations (8) and (9) to the actual data simultaneously. Fitting was done using the *nlinfit* function available in the MATLAB R2019b [17] software. 95% confidence intervals for the predicted values are obtained by using the *nlpredci* function. 95% confidence interval for the fitted parameters *I*(0), *a, h* and *µ* are found using the *nlparci* function.

## Results

We first tested our model on the data of the Bombay Plague Epidemic, 1905-6. In [2], Kermack and McKendrick fitted the data of deaths to obtain a model defined by the function 890 *sech*^2^(0.2t - 3.4), where *t* represents time in weeks. Since we didn’t get real data, we used the above function to model the active cases. The total cases reported were also computed by taking cumulative sum using the above function. This could be justified as most of the patients who contracted the disease had died. Figure 1 shows the fit (together with the 95% confidence intervals) for the active and the total cases by considering them as generated by quations (8) and (9) simultaneously. The figure shows that our combined model produced a nice fit of the above data. We then fitted the active cases data using equation (8) alone and the total cases data using equation (9); we obtained a nice fit in both cases. Table 1 shows the fitted parameters. Though this is the case, the parameters obtained by fitting active cases using equation (8) alone produced a poor fit of the total cases curve and vice versa. This persuaded us to rely on the combined model rather than on the individual models. Bacaer [19], which studied the Kermack and McKendrick model based on the Bombay epidemic data, discusses about the possibility of variation in the basic reproduction number and gives sample values for *R*_0_ as 1.09, 1.17 and 1.24 (please refer Table 1 in [19]). Figure 2 shows the movement of the time dependent basic reproduction number *R*_0_(*t*), found using equation (12), through the infection period. For the combined model, it started from 2.39 and then decreased whereas in the active cases individual model it started from 1.43 before decreasing. The stationary point *t*_*s*_ computed by the combined model, active cases individual model and total cases individual model is 17.06, 16.92 and 16.76 respectively; whereas the Kermack and McKendrick model gives it as 17.

**Table 1.** Parameter values for the fitted model to the data of the Bombay Plague Epidemic, 1905-6 [2]. We have considered three cases. The first one is the evaluation of the parameters by considering equations (8) and (9) simultaneously; second using equation (8) for the actives cases and third using equation (9) for total cases reported.

| Parameter | | a | b | $\mu$ | $I(0)$ |
| --- | --- | --- | --- | --- | --- |
| Equations (8) and (9) simultaneously | Value | 2.5561 | 0.0542 | 1.0132 | 0.0116 |
|  | 95% Confidence Bounds | (2.46, 2.65) | (0.051, 0.057) | (0.96, 1.07) | (0.005, 0.018) |
| Equation (8) alone | Value | 4.0911 | 0.0226 | 2.7913 | 0.0292 |
|  | 95% Confidence Bounds | (0.57, 7.61) | (-0.009, 0.05) | (-1.09, 6.68) | (-0.03, 0.08) |
| Equation (9) alone | Value | 3.2728 | 0.0273 | 2.0699 | 0.0366 |
|  | 95% Confidence Bounds | (1.01, 5.53) | (-0.006, 0.061) | (-0.53, 4.68) | (0.006, 0.067) |

**Figure 1.**
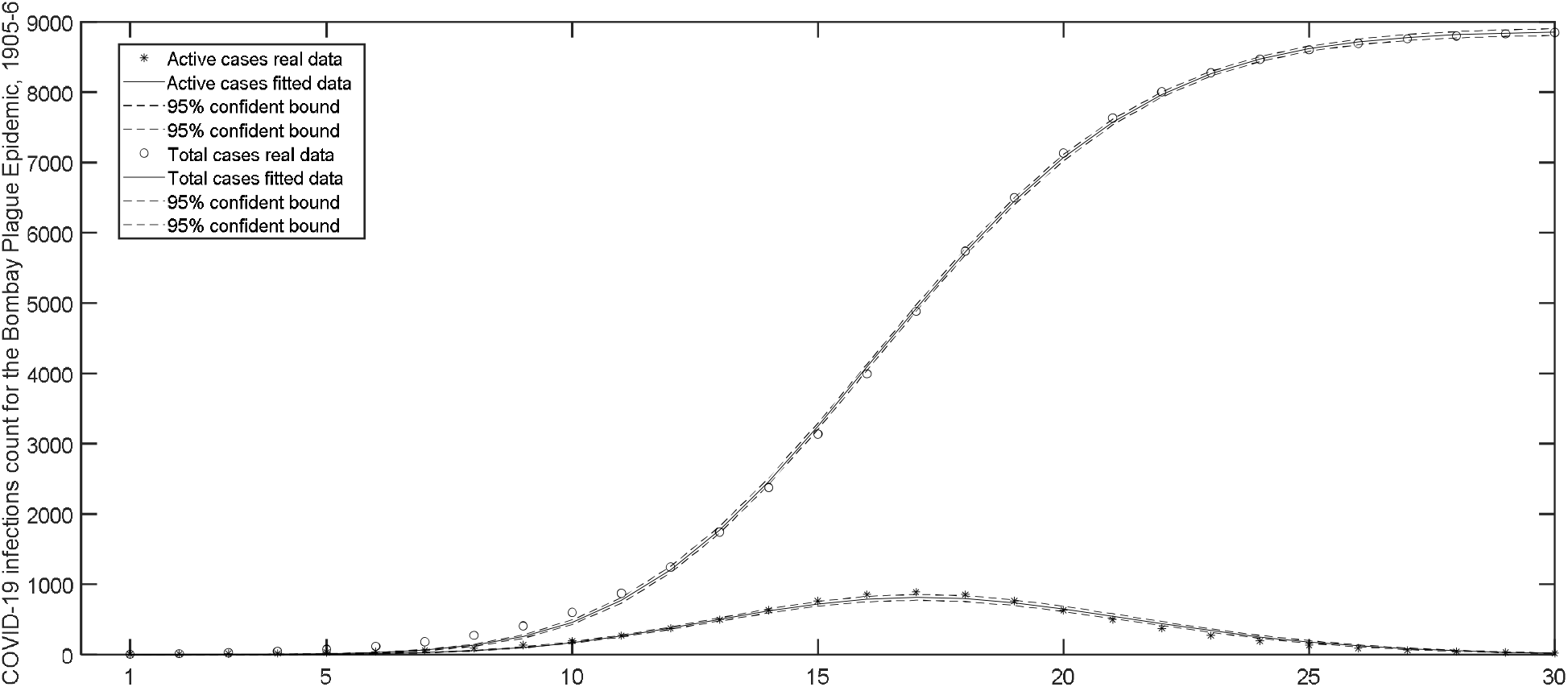
Fit of our model to the data of the Bombay Plague Epidemic, 1905-6 [2] by considering equations (8) and (9) simultaneously.

**Figure 2.**
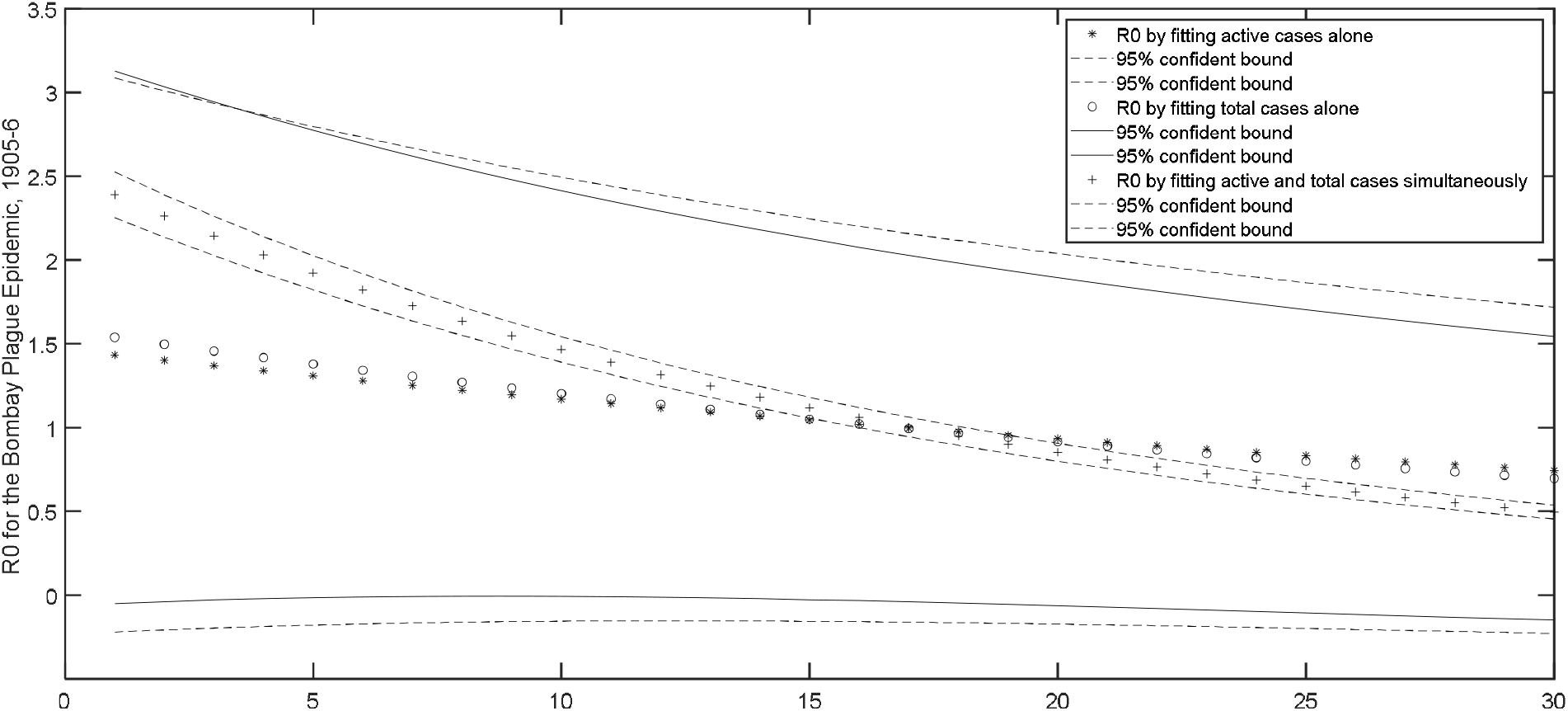
Behavior of *R*_0_(*t*) for the data of the Bombay Plague Epidemic, 1905-6 [2], when it is calculated on parameters obtained by: (i) fitting active cases using equation (8) alone (ii) fitting total cases using equation (9) alone (iii) fitting active and total cases using equations (8) and (9) simultaneously.

Now we turn our attention to the COVID-19 data. We fitted the model defined by equations (8) and (9) to the data showing the number of COVID-19 infected people (active cases and total reported cases) in several countries from January 22 to April 10, 2020. We first fitted our model to the data for the Mainland China from January 22 to April 10, 2020 [15]. Figure 3 shows the fit of the active and the total cases by applying the combined model. The figure shows that the fit misses the data. Suspecting this may be due to fitting by the combined model, we fitted these equations separately. This means we assume that the active cases data is generated by equation (8) alone and the total cases by equation (9). The resulting figure, Figure 4 shows a good fit for both active and total cases. The fitted parameter values for simultaneous and other fits are given in Table 2.

**Table 2.** Parameter values for the fitted model to the data showing the daily count of COVID-19 infected people in China [15] from January 22 to April 10, 2020. We have considered three cases. The first one is the evaluation of the parameters by considering equations (8) and (9) simultaneously; second using equation (8) for the actives cases and third using equation (9) for total COVID-19 cases reported.

| Parameter | | a | b | $\mu$ | $I(0)$ |
| --- | --- | --- | --- | --- | --- |
| Equations (8) and (9) simultaneously | Value | 2.2966 | 0.1536 | 0.0483 | 0.0685 |
|  | 95% Confidence Bounds | (1.43, 3.17) | (0.14, 0.17) | (0.04, 0.05) | (-0.2, 0.34) |
| Equation (8) alone | Value | 0.6743 | 0.0583 | 0.1367 | 233.2053 |
|  | 95% Confidence Bounds | (0.6, 0.74) | (0.049, 0.07) | (0.12, 0.16) | (97.5, 368.9) |
| Equation (9) alone | Value | 1.2788 | 0.0187 | 0.8981 | 157.7497 |
|  | 95% Confidence Bounds | (-0.47, 3.02) | (-0.02, 0.06) | (-1.0, 2.8) | (26.3, 289.2) |

**Figure 3.**
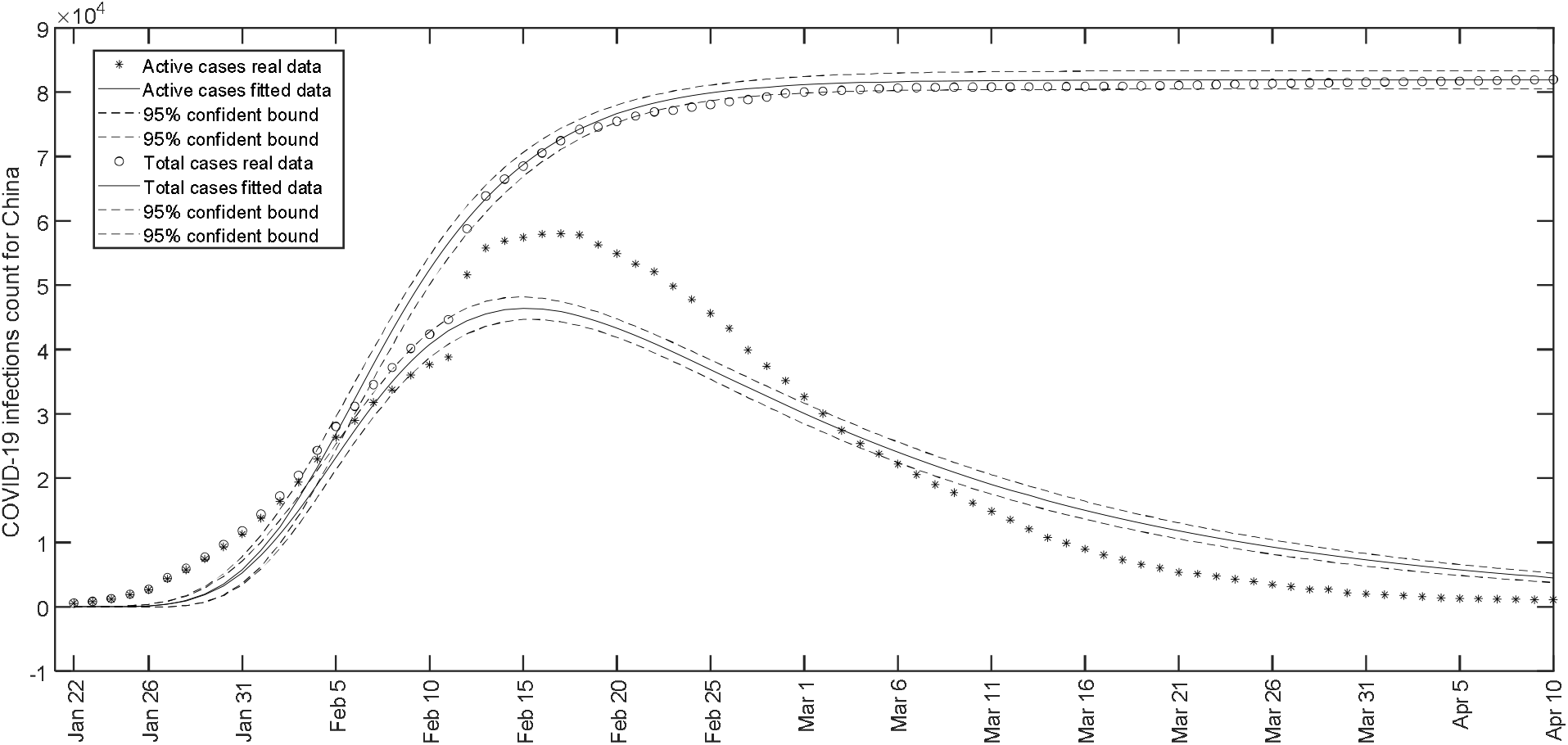
Fit of the model to the data showing the count of COVID-19 active and total cases in China [15] by considering equations (8) and (9) simultaneously. Day 1 stands for January 22 and Day 80 for April 10, 2020.

**Figure 4.**
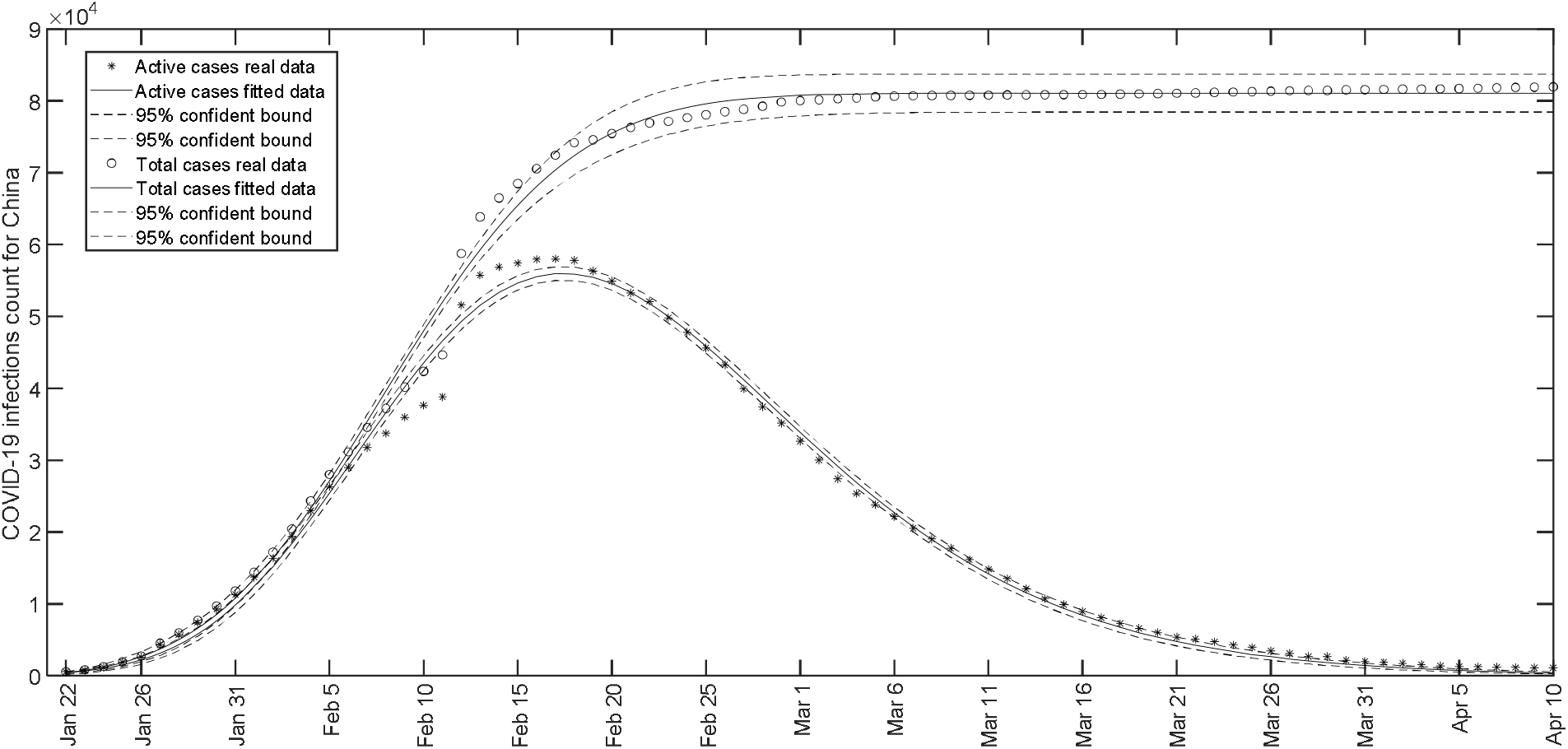
Fit of the model to the data showing the count of COVID-19 active cases (using equation (8) alone) and total cases (using equation (9) alone) in China [15].

The stationary point *t*_*s*_, calculated from the parameters obtained by fitting active cases using equation (8) alone, is day 27.4. Since Day 1 is January 22, Day 27 turns out to be February 17. The active cases graph for China [15] shows that the number of cases did stop increasing on February 17. However when parameters from the combined model are considered, we obtained *t*_*s*_ as 25.2. Data reveals that there was a drastic increase in the active number of cases on February 12 to 51591 from 38791 on the previous day. Variations like this and much severe can be observed during the COVID-19 progression for many countries. This makes the future prediction, based on a set of fitted parameters at a particular time point, very difficult. This demands updating the fit from time to time. We have found that the combined model would have yielded *t*_*s*_ as: 32.03 for fit up to February 11; 49.4 up to February 13; 41.2 up to February 17; 33.6 up to February 21; 29.1 up to February 27.

Figure 5 shows the movement of the basic reproduction number during the period starting from January 22 to April 10, 2020. This is obtained by plotting *R*_0_(*t*) given in equation (12) taking the parameters shown in Table 2 by varying time *t* from 1 (January 22) to 80 (April 10). For the model based on equation (8) alone *R*_0_(*t*) started from 4.65; whereas in the case of model based on equation (9) alone it started from 1.4 for day 1 and then decreased. These estimates agree with other studies in this direction [18]. However for the combined model, *R*_0_(*t*) started from a large value 40.8 on January 22, dropped to 10.2 on January 31, became 4.7 on February 5 and then decreased. We would have abandoned these results from the joint model considering the mismatch with data (please see Figure 3), if this wasn’t repeated for Italy also.

**Figure 5.**
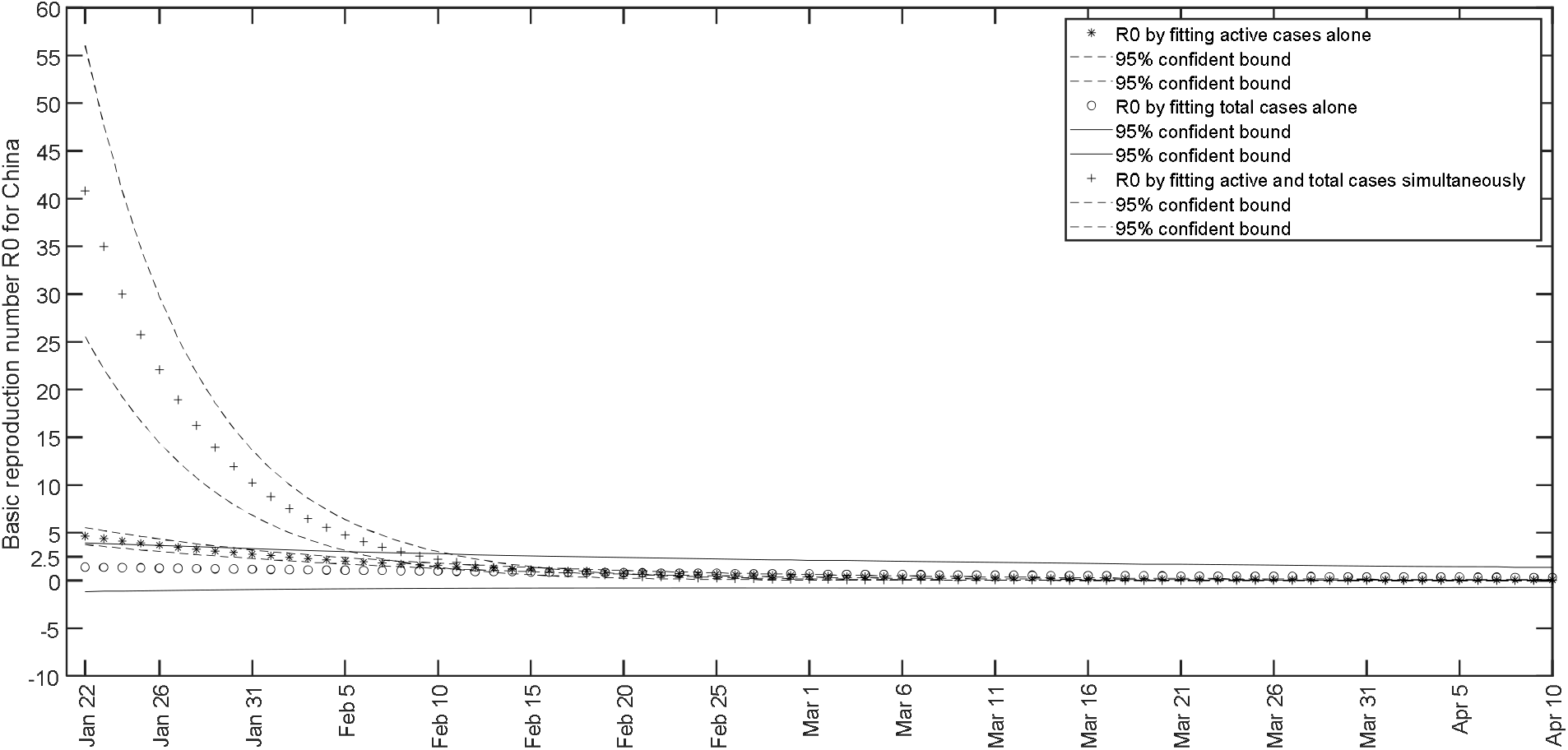
Behavior of *R*_0_(*t*) for China from January 22 to April 10, 2020 when it is calculated on parameters obtained by: (i) fitting active cases using equation (8) alone (ii) fitting total cases using equation (9) alone (iii) fitting active and total cases using equations (8) and (9) simultaneously.

For Italy we fitted the data from February 15 (Day 1) to April 10 (Day 56), 2020. Unlike in the case of China, here the combined model produced a good fit of the data. The fit is given in Figure 6. The parameter estimates produced by the combined and the individual models are given in Table 3. On fitting the combined model to the data up to March 21, 2020, we obtained the parameters as a=0.4823, b=0.0331, *µ*=0.0363 and *N*(0)=6.1616. The stationary point *t*_*s*_ was obtained as 78.1. However for the fit up to March 25, the stationary point *t*_*s*_ decreased to 64.8. The parameters for this fit are a=0.6911, b=0.0458, *µ*=0.0355 and *N*(0)=0.7589. Hence our model suggests a decrease in the disease spreading rate in Italy by March 25. *t*_*s*_ moved to 59.4 for a fit up to March 31; to 58.5 up to April 5, 2020 and became 59.1 on fitting data up to April 10.Though this may be signaling a decrease in the infection rate between March 25 and April 10, it may also be due to an increase in the patient death rate than the recovery rate. To check this we fitted the combined model governed by equations (8) and (9) to the data of the (active + dead) cases and the total cases. This is equivalent to assume that a decrease in the number of infected cases occurs due to patient recovery only. For *t*_*s*_, We obtained 90.7 in place of 78.1 by fitting the data up to March 21; 73.97 in place of 64.8 up to March 25; 66.8 in place of 59.4 up to March 31; 65.0 in place of 58.5 up to April 5 and 65.03 in place of 59.1 for a fit up to April 10, 2020. This again shows that the infection is certainly suppressing in Italy and if the pattern continued, the infection count may start decreasing from the second or third week of April, 2020.

**Table 3.**
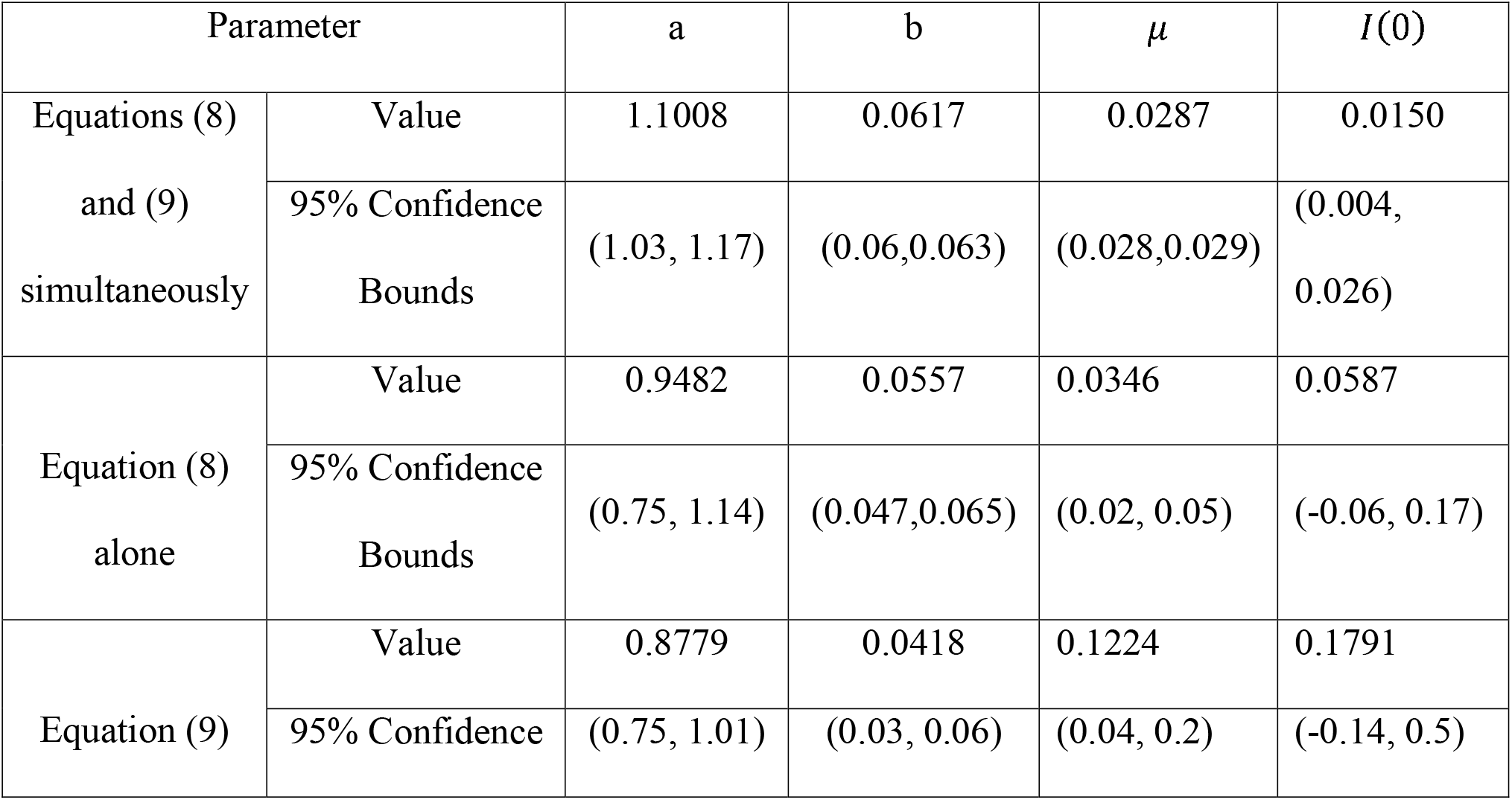
Parameter values for the fitted model to the data showing the daily count of COVID-19 infected people in Italy [16] from February 15 to April 10, 2020. We considered three cases. The first one is the evaluation of the parameters by considering equations (8) and (9) simultaneously; second using equation (8) for the actives cases and third using equation (9) for total COVID-19 cases reported.

**Figure 6.**
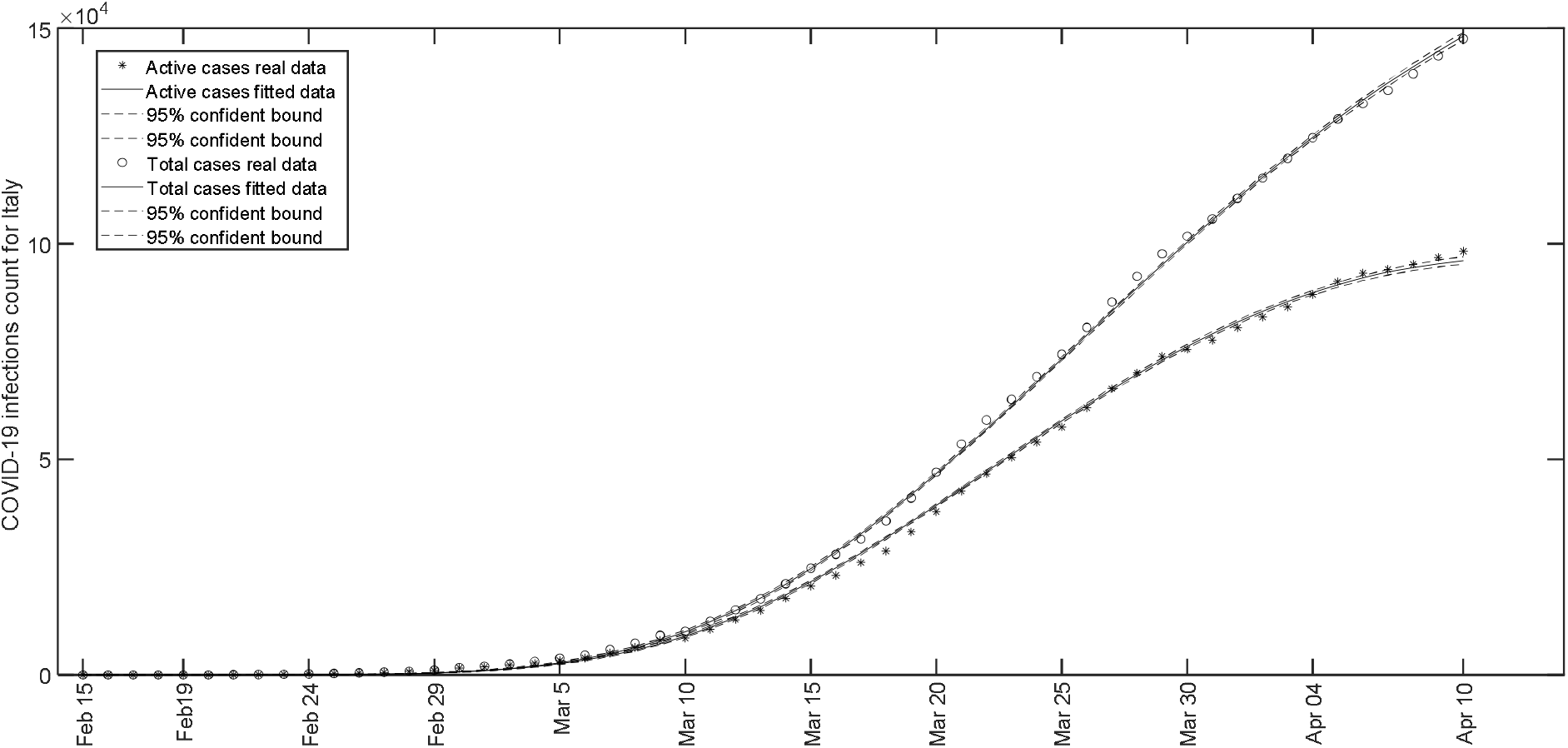
Fit of the model to the data showing the count of COVID-19 active and total cases in Italy [16] by considering equations (8) and (9) simultaneously. Day 1 stands for February 15 and Day 56 for April 10, 2020.

Figure 7 shows that a similar fit as in the case of the combined model has been obtained in the case of individual fits of equations (8) and (9). However, the parameter estimation using individual models, either (8) or (9), can’t be considered reliable always. For example, a fit to the data up to March 21 produced negative values for the parameter *µ* by considering both individual models (8) and (9).

**Figure 7.**
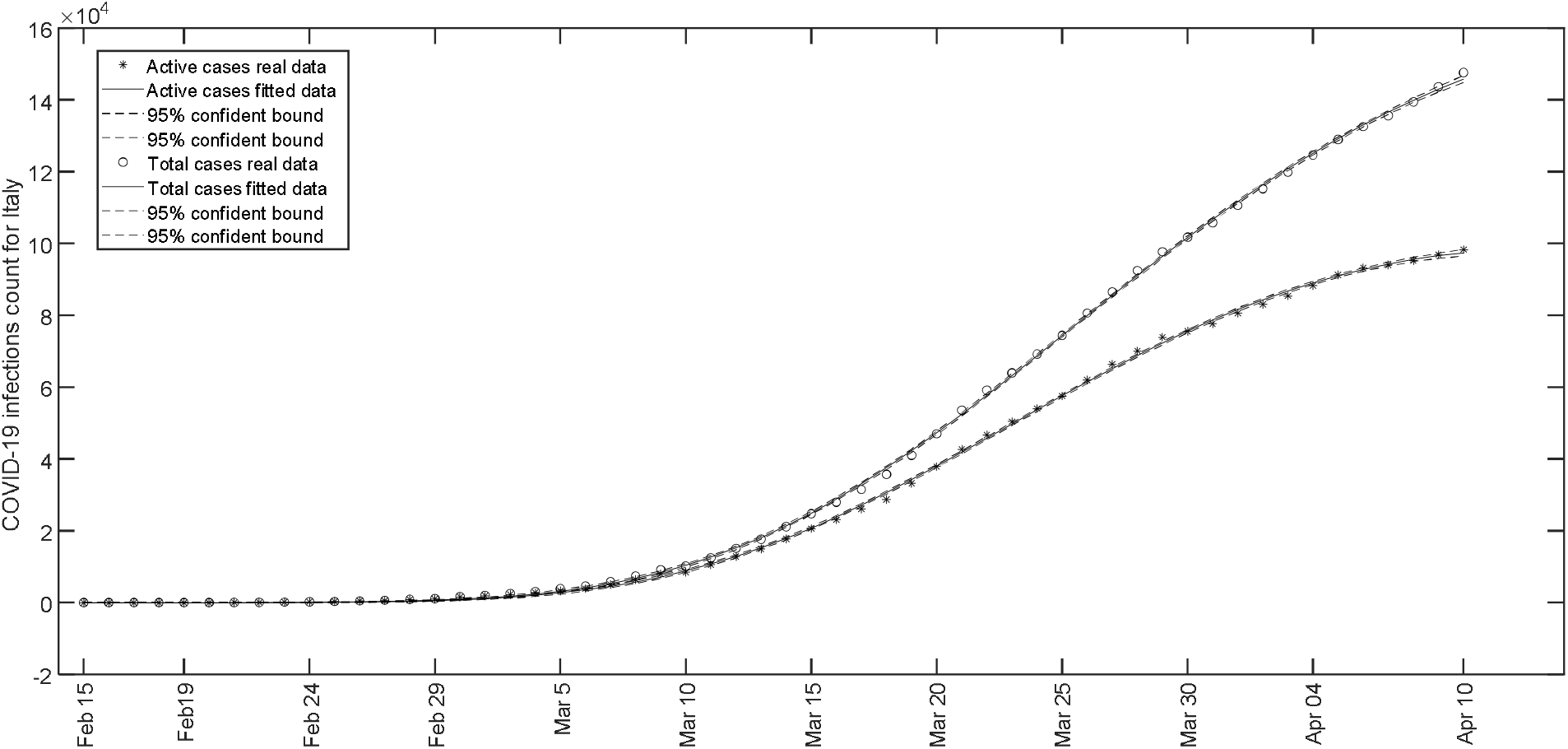
Fit of the model to the data showing the count of COVID-19 active cases (using equation (8) alone) and total cases (using equation (9) alone) in Italy [16].

Figure 8 show the movement of the function *R*_0_(*t*) from February 15 to April 10, 2020. In the active, total and combined model cases it started from 25.9, 6.9 and 36.1 respectively and then decreased. After 40 days, that is on March 25, these values were respectively 2.95, 1.3 and 3.25. On April 10, 2020 these values became 1.21, 0.69 and 1.21. According to the current fit, it shall become 0.73, 0.47 and 0.70 on April 19, 2020.

**Figure 8.**
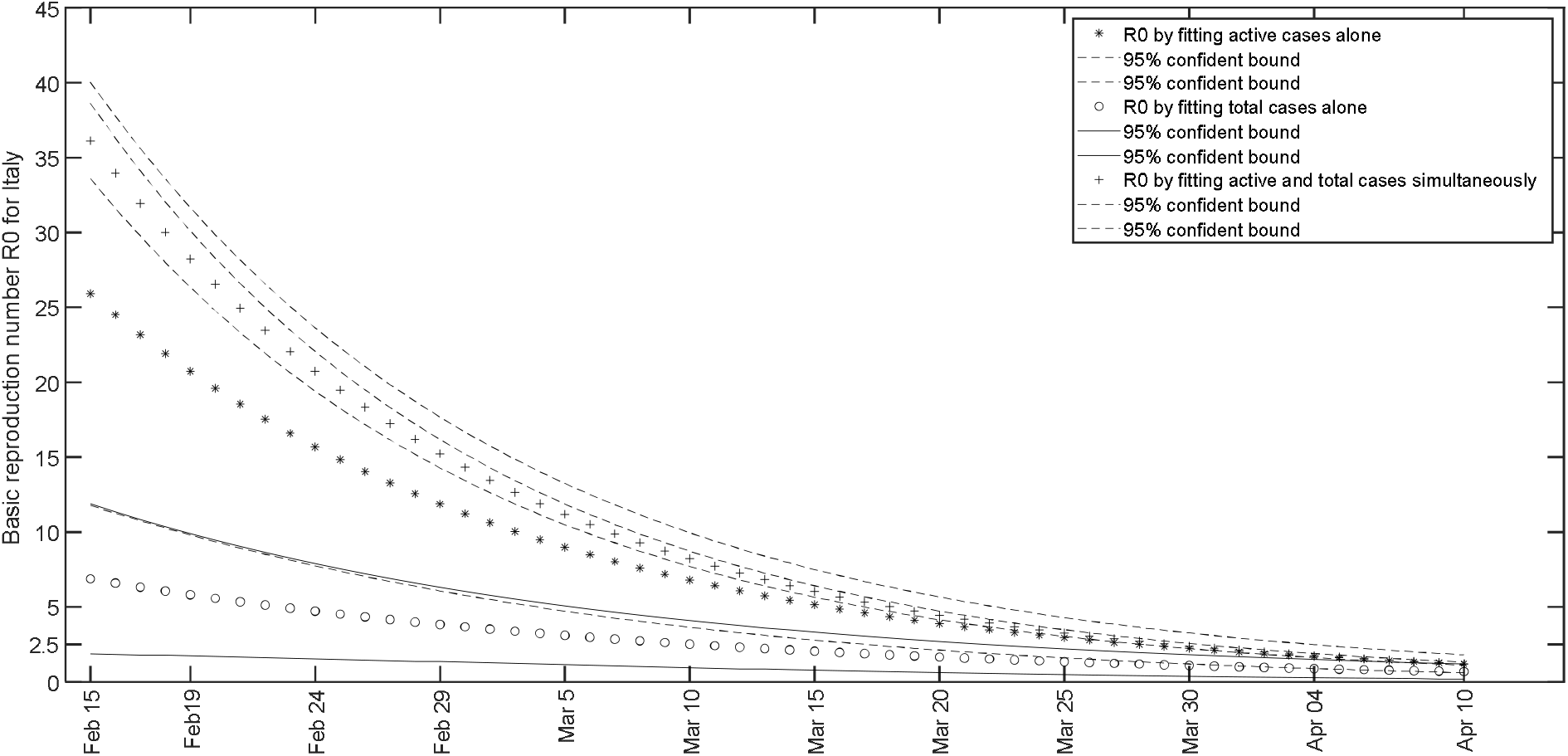
Behavior of *R*_0_(*t*) for Italy from February 15 to April 10, 2020 when it is calculated on parameters obtained by: (i) fitting active cases using equation (8) alone (ii) fitting total cases using equation (9) alone (iii) fitting active and total cases using equations (8) and (9) simultaneously.

Model fits to the data for the countries: India, United States, Germany and Canada are given in the annexure.

## Conclusion

We have developed a birth and death model for fitting the COVID-19 active/(active + dead) and total cases. Our model turns out to be a special case of the standard SIR model. The advantage of our model is its analytic form, which is suitable for fitting the data. As a result, important characteristics of the disease progression path can be studied. We have seen that the model fits the data for several countries. However the fit needs to be updated for more realistic future prediction. This means that the parameter estimates given in the paper may not be of use for some countries as time elapses. Please refer the annexure to view the effect of parameter update on the future prediction.

## Data Availability

The COVID-19 cases data can be found in the website https://www.worldometers.info.

## Declarations

### Availability of data and material

The COVID-19 cases data can be found in the website https://www.worldometers.info.

### Code availability

The MATLAB code used for fitting the model can be obtained from the author N C Viswanath upon request.

## Annexure

### Analysis and Prediction of COVID-19 Characteristics Using a Birth-and-Death Model

In the case of Italy, we obtained a better future prediction when we considered the combined model rather than the individual models; also we had a better prediction of the stationary point when we considered active + dead cases instead of just active cases. Therefore we present the results obtained by fitting the combined model defined by the set of equations (8) and (9) to the COVID-19 data of active + dead cases and total cases in different countries between February 15 and June 15, 2020.

### India

In the case of India, a fit (active + dead and total cases) from February 15 up to June 15 revealed t_S_ as equal to 183.4 that is 16^th^ August 2020. The fit is given in Figure 1 and the parameters are given in Table1. The basic reproduction number *R*_0_(*t*) started from 4.54 on Day 1 (February 15) and decreased to become 0.995 on Day 184 (August 16); Figure 2 shows its movement. On fitting the active and total cases we found *t*_*S*_ as 179.9, *R*_0_(1) as 4.15 and *R*_0_(184) as 0.97.

**Table 1.**
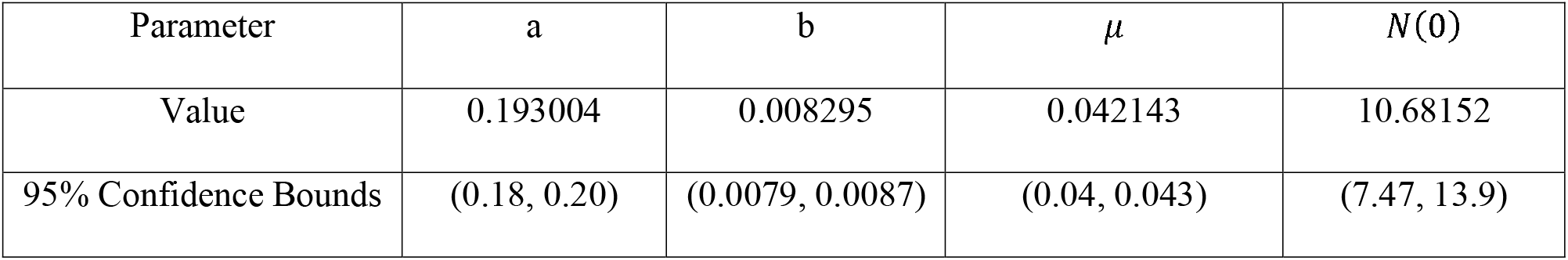
Parameter values for the fitted combined model to the data showing the daily count of COVID-19 active + dead cases and total cases from February 15 to June 15, 2020 in India^1^.

| Parameter | a | b | $\mu$ | $N(0)$ |
| --- | --- | --- | --- | --- |
| Value | 0.193004 | 0.008295 | 0.042143 | 10.68152 |
| 95% Confidence Bounds | (0.18, 0.20) | (0.0079, 0.0087) | (0.04, 0.043) | (7.47, 13.9) |

**Figure 1.**
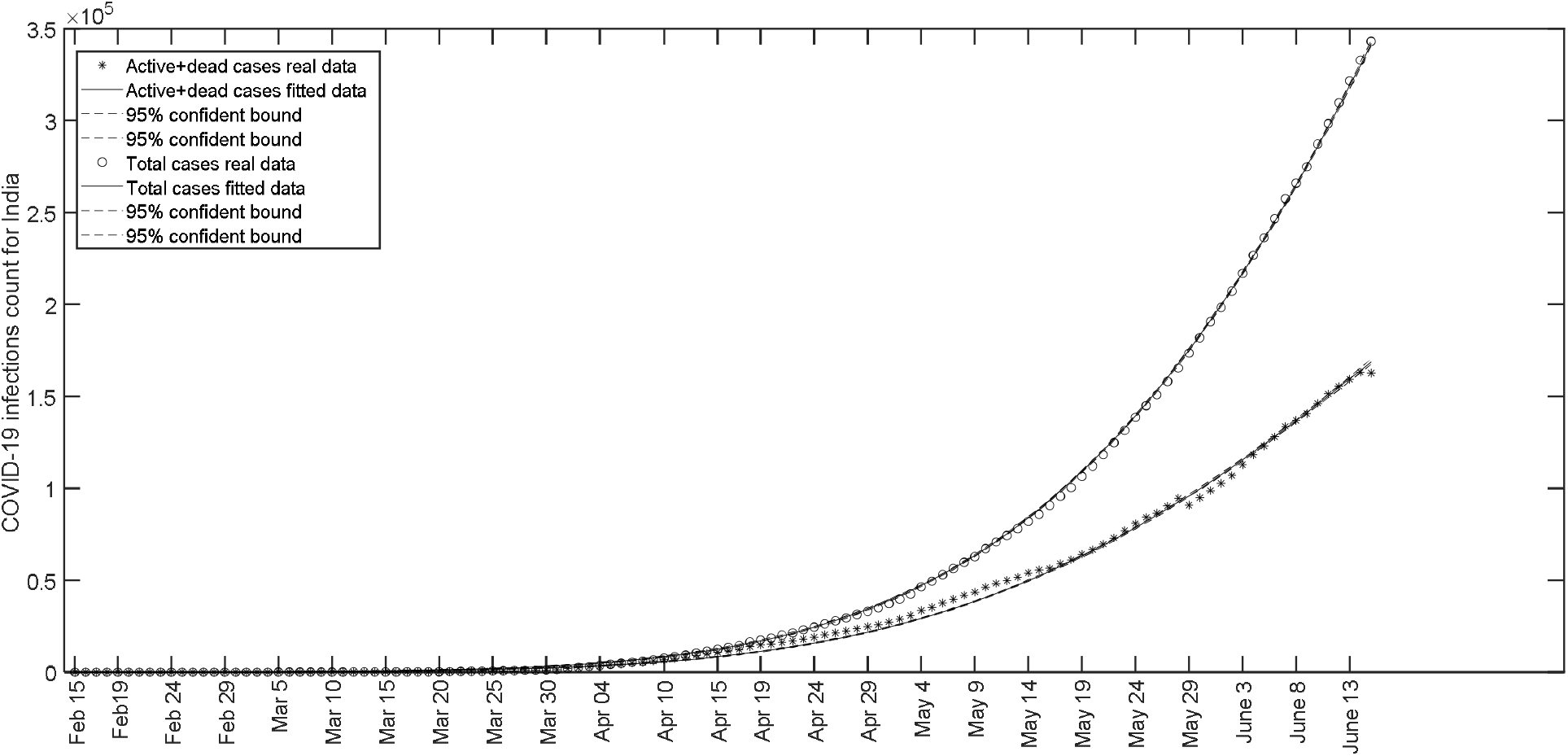
Fit of the model to the data showing the daily count of COVID-19 active + dead cases and total cases from February 15 to June 15, 2020 in India^1^.

**Figure 2.**
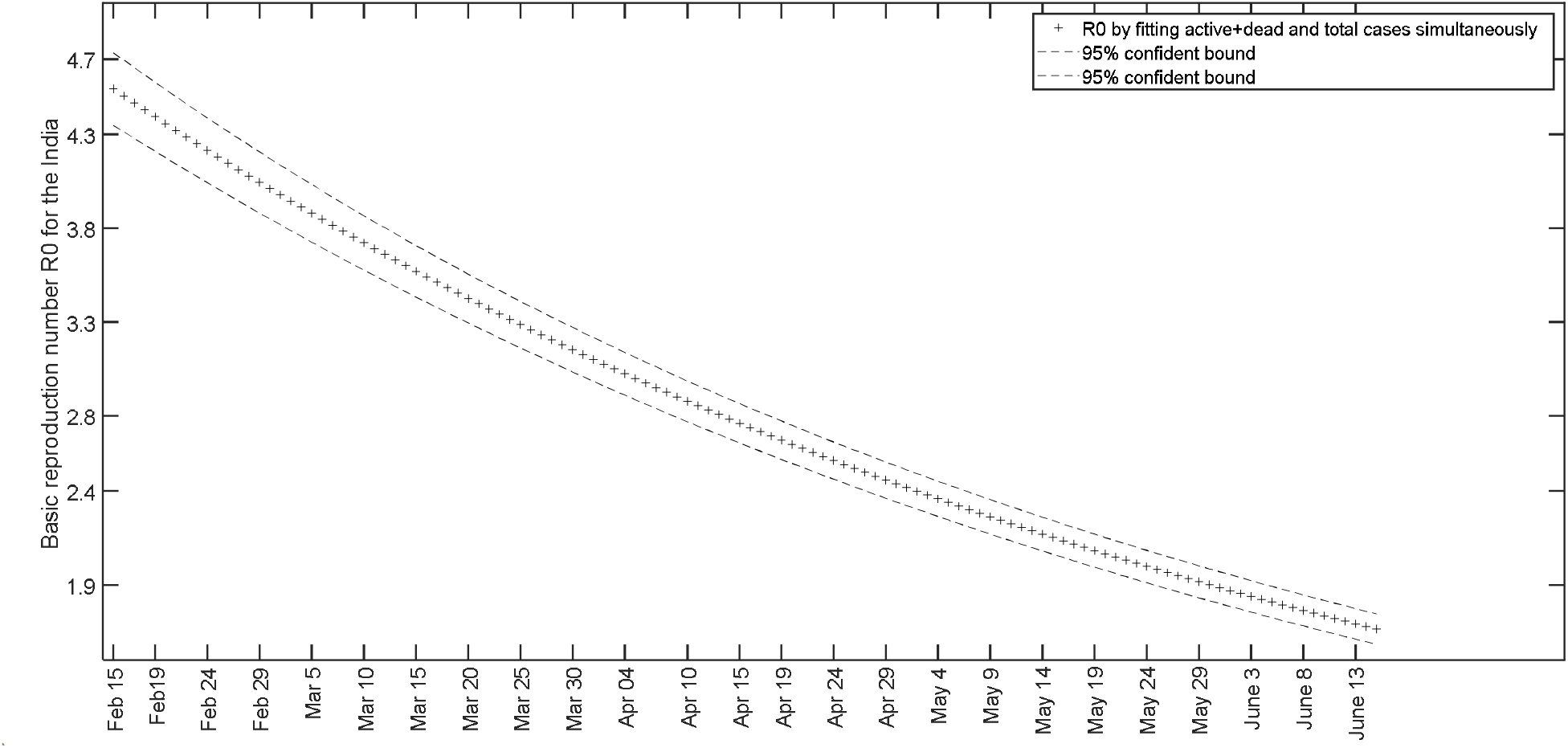
Behavior of *R*_*0*_(*t*)for India from February 15 to June 15, 2020 when it is calculated on parameters obtained by fitting active + dead and total cases using equations (8) and (9) simultaneously

### The United States of America

In the case of The United States of America, a fit (active + dead and total cases) from February 15 up to June 15 revealed t_S_ as equal to 111.2 that is June 5. The fit is given in Figure 3 and the parameters are given in Table 2. The basic reproduction number *R*_0_(*t*) started from 35.7 on Day 1 (February 15) and decreased to become 0.97 on Day 112 (June 5); Figure 4 shows its movement. On fitting the active and total cases we found *t*_*S*_ as 106.98, *R*_0_(1) as 28.02 and *R*_0_(107) as 0.999.

**Table 2.** Parameter values for the fitted combined model to the data showing the daily count of COVID-19 active + dead cases and total cases from February 15 to June 15, 2020 in The United States of America

| Parameter | a | b | $\mu$ | $N(0)$ |
| --- | --- | --- | --- | --- |
| Value | 0.410585 | 0.032434 | 0.011138 | 19.7158 |
| 95% Confidence Bounds | (0.38, 0.44) | (0.031, 0.034) | (0.01, 0.0115) | (6.28, 33.16) |

**Figure 3.**
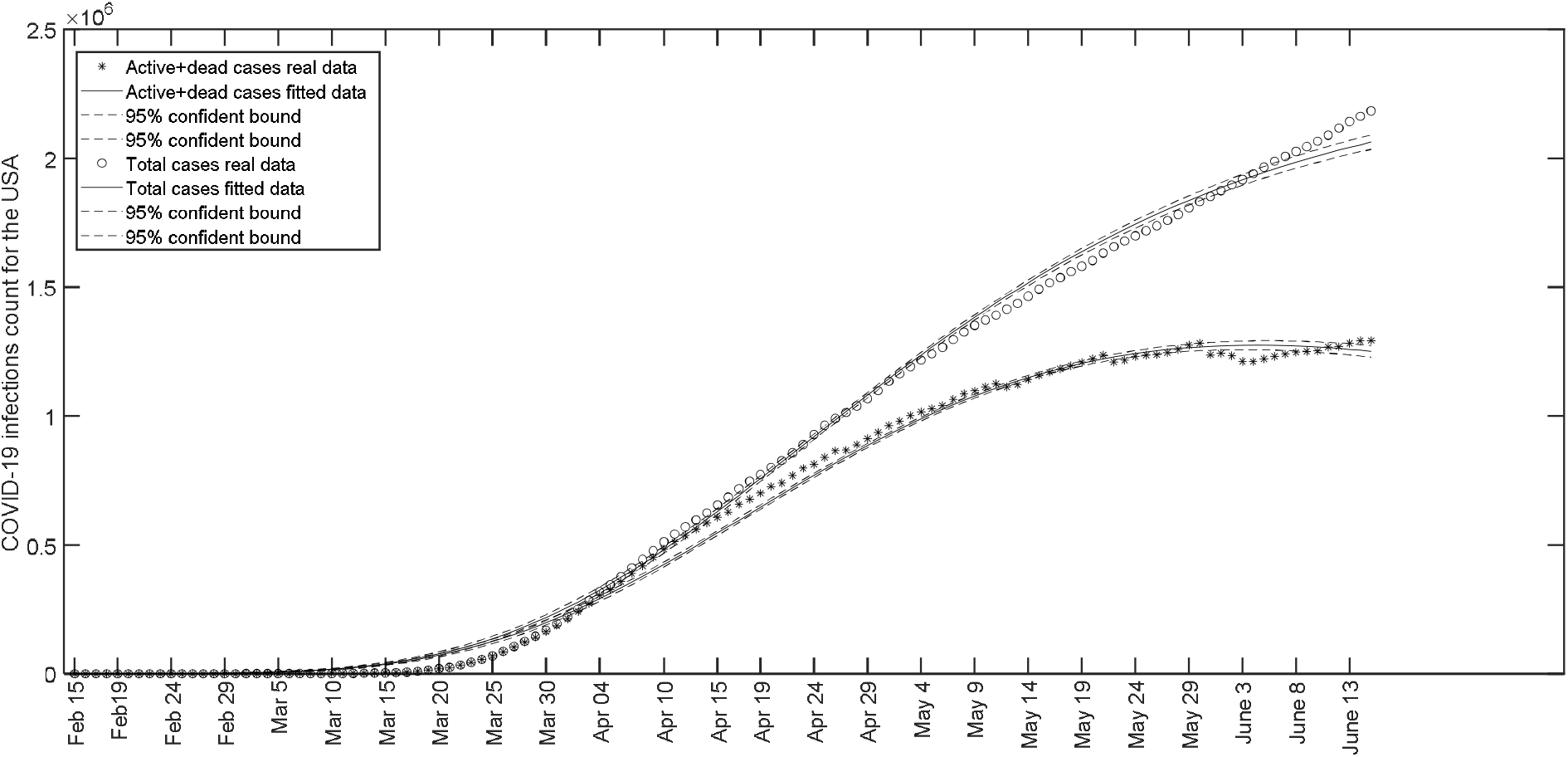
Fit of the model to the data showing the daily count of COVID-19 active + dead cases and total cases from February 15 to June 15, 2020 in The United States of America^2^.

**Figure 4.**
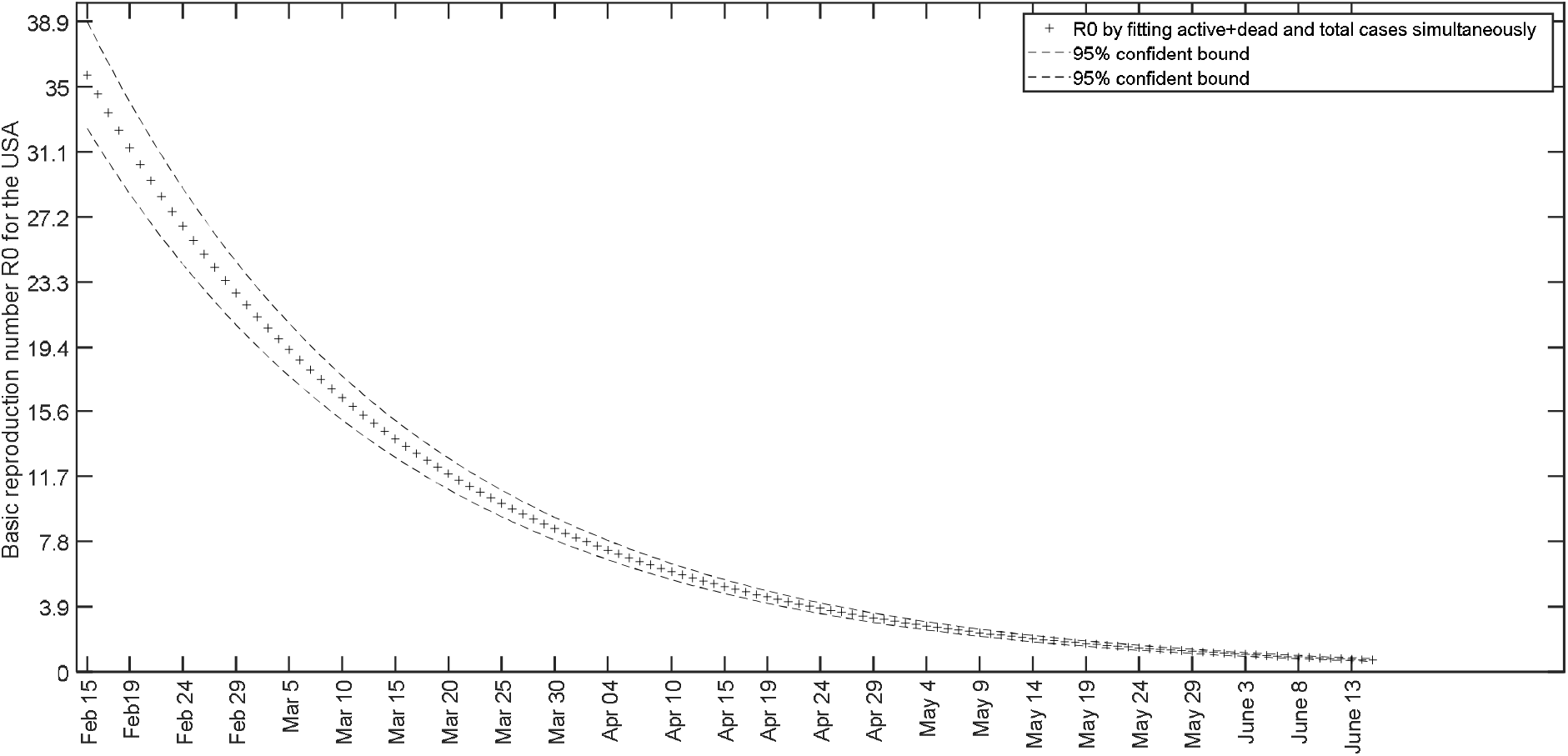
Behavior of *R*_*0*_(*t*) for The United States of America from February 15 to June 15, 2020 when it is calculated on parameters obtained by fitting active + dead and total cases using equations (8) and (9) simultaneously.

### Germany

In the case of Germany, the fit was performed from February 15 up to June 15. It revealed t_S_ as equal to 58.7; that is April 13, 2020. The fit is given in Figure 5 and the parameters are given in Table 3. The basic reproduction number *R*_0_(*t*) started from 20.3 on Day 1 (February 15) and decreased to 0.98 on Day 59 (April 13). The movement of *R*_0_(*t*) is shown in Figure 6. A fit of active and total cases revealed *t*_*S*_ as 57.2, R_0_(1) as 19.4 and *R*_0_ (59) as 0.91. This shows that there is not much difference between active cases fit and active + dead cases fit for Germany. High recovery as well low death rate in Germany could be the reason for this phenomenon.

**Table 3.**
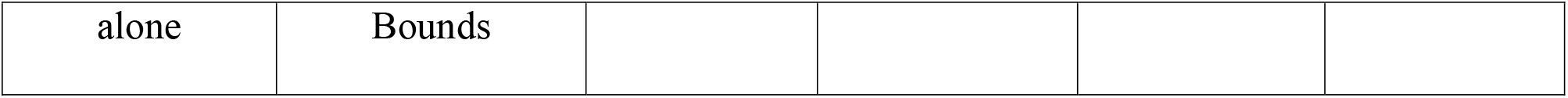
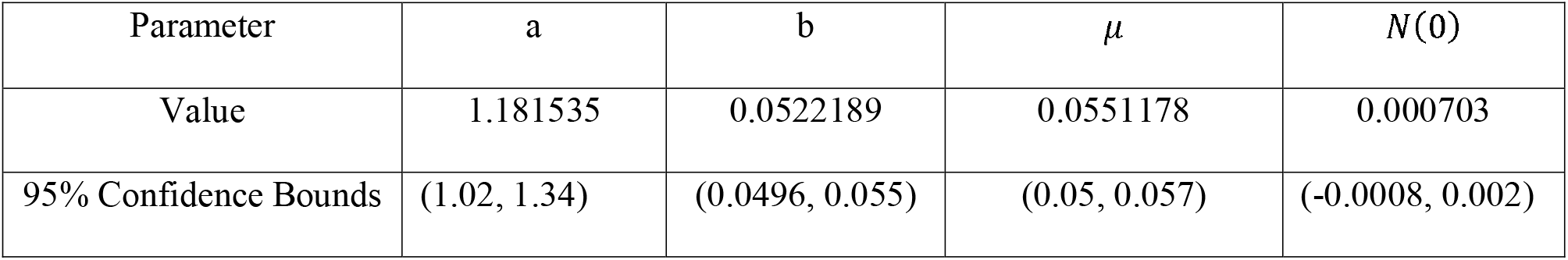
Parameter values for the fitted combined model to the data showing the daily count of COVID-19 active + dead cases and total cases from February 15 up to June 15 in Germany^3^.

**Figure 5.**
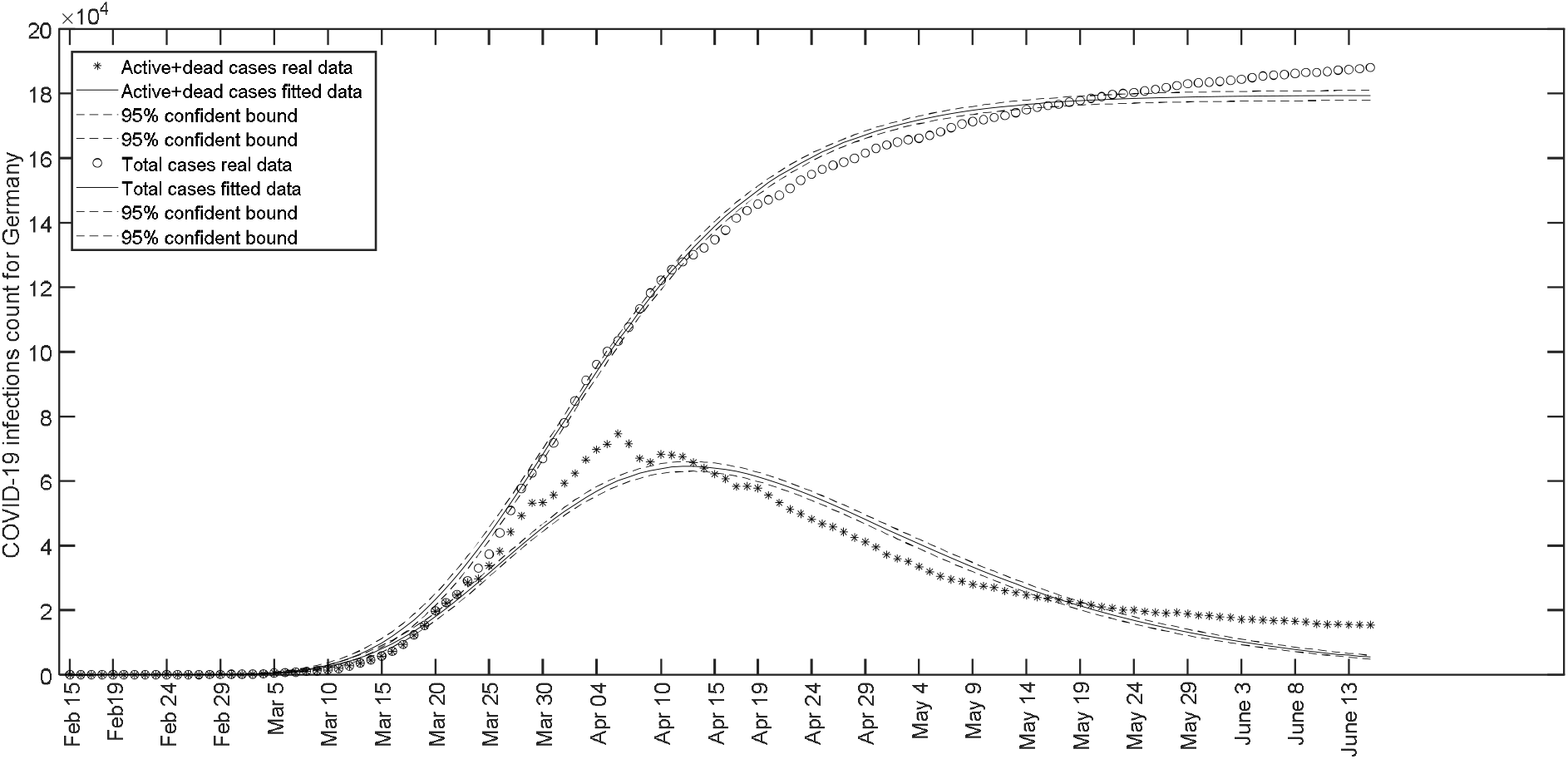
Fit of the model to the data showing the daily count of COVID-19 active + dead cases and total cases from February 15 up to June 15 in Germany^3^.

**Figure 6.**
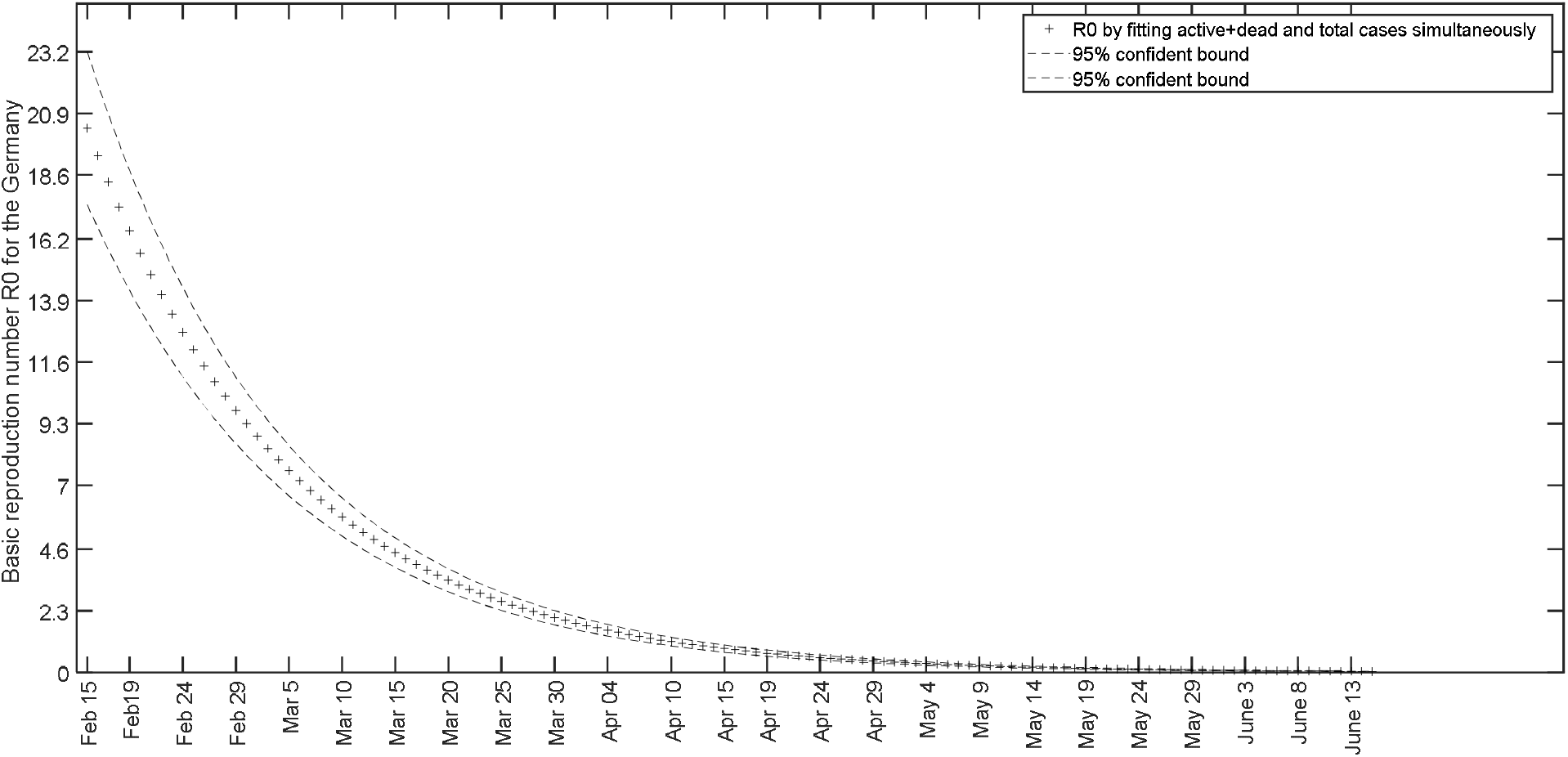
Behavior of *R*_*0*_(*t*) for Germany from February 15 to June 15, 2020 when it is calculated on parameters obtained by fitting active + dead and total cases using equations (8) and (9) simultaneously.

### Canada

In the case of Canada, a fit February 15 up to June 15. It revealed t_S_to be 98.2 that is May 23, 2020. The fit is given in Figure 7 and the parameters are given in Table 4. The basic reproduction number R_0_(t) started from 13.96 on Day 1 (February 15) and decreased to 0.98 on Day 99 (May 23). The movement of R_0_(t) is shown in Figure 8. A fit of active and total cases revealed t_S_ as 94.1, R_0_(1) as 10.3 and R_0_(99) as 0.88.

**Table 4.** Parameter values for the fitted combined model to the data showing the daily count of COVID-19 active + dead cases and total cases in Canada^4^ from February 15 up to June 15.

| Parameter | a | b | $\mu$ | $N(0)$ |
| --- | --- | --- | --- | --- |
| Value | 0.375898 | 0.0271191 | 0.0262127 | 1.37559 |
| 95% Confidence Bounds | (0.35, 0.4) | (0.026, 0.028) | (0.0257, 0.0267) | (0.66, 2.1) |

**Figure 7.**
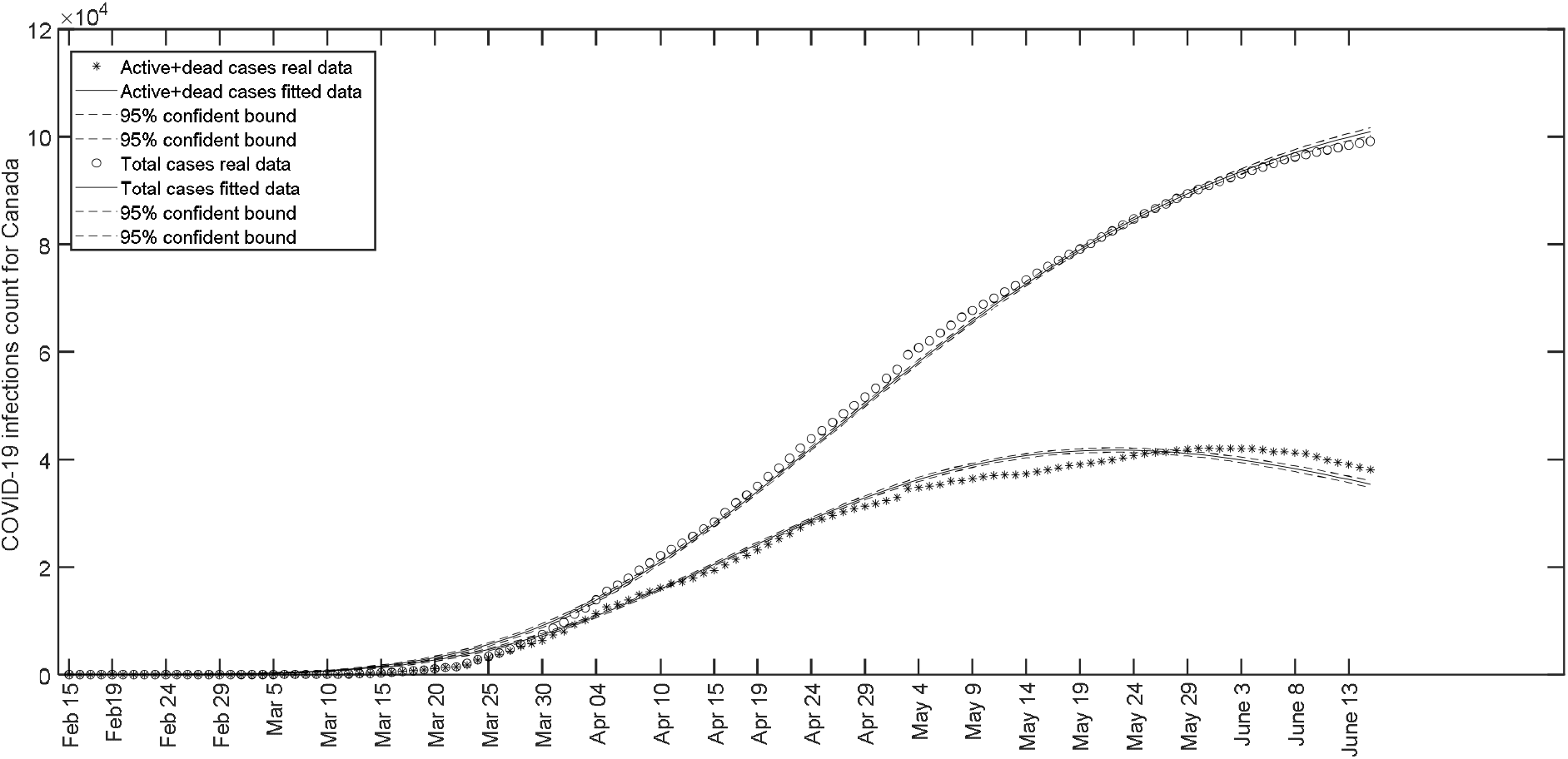
Fit of the model to the data showing the daily count of COVID-19 active + dead cases and total cases in Canada4 from February 15 up to June 15.

**Figure 8.**
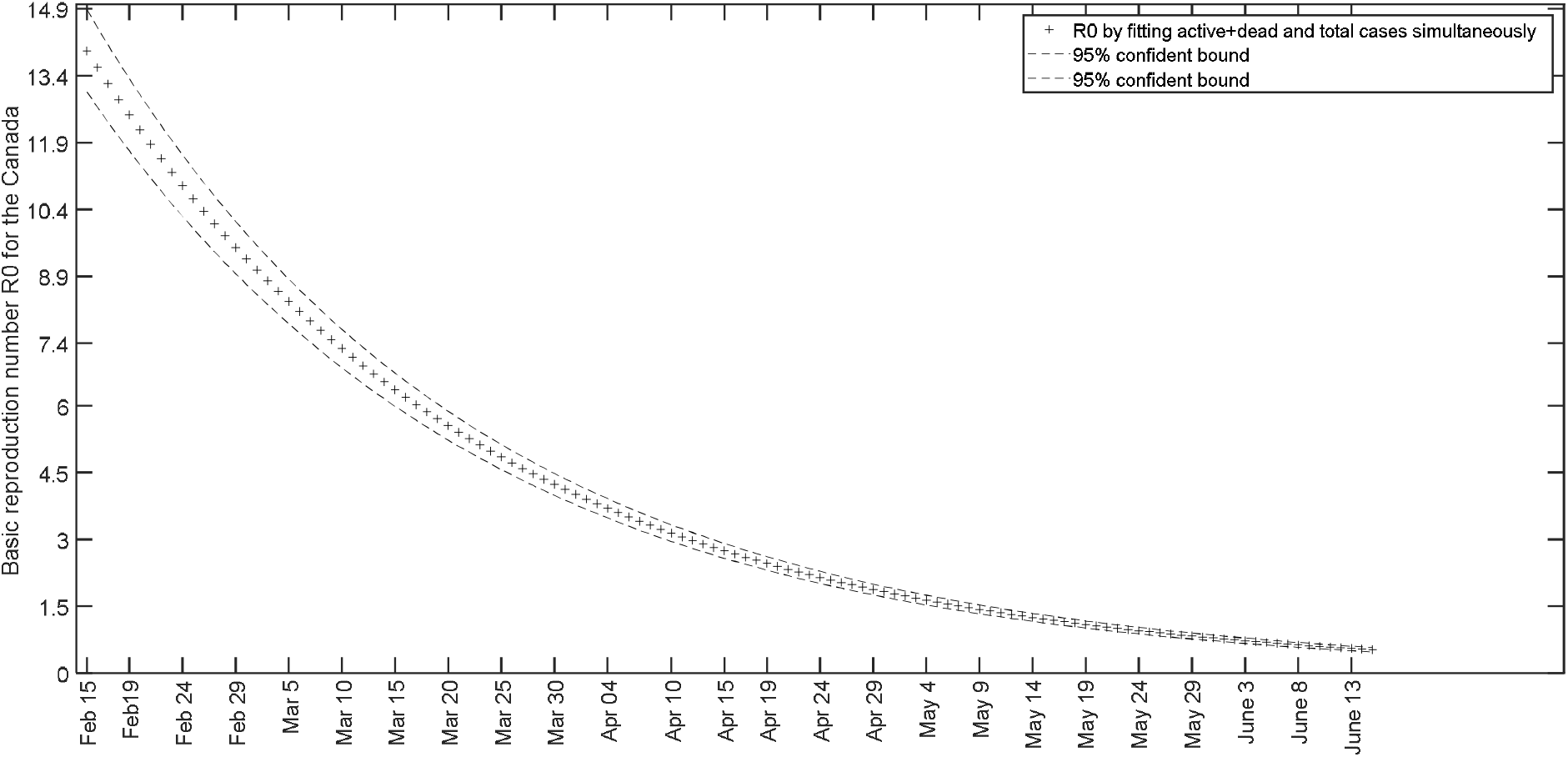
Behavior of *R*_*0*_(*t*) for Canada from February 15 up to June 15 when it is calculated on parameters obtained by fitting active + dead and total cases using equations (8) and (9) simultaneously.

### Comparison of future prediction based on data for various periods

Here we discuss what could have been the predictions by our model for a future date, if it is fitted against the data up to certain date in the past. More precisely, we compare the values predicted by our model with the actual values for June 15 based on data up to different past dates.

Table 5 shows the predictions based on the data up to April 20, 2020. It shows that except for Italy and Germany, where the active cases started decreasing before April 20, the predictions differed from the actual values. Table 6 shows the prediction based on the data up to May 20, 2020. It shows that better predictions are obtained with increase in data. However the predictions varied largely from actual data for the countries India and USA, where the disease showed faster progress near June 20. When we extended the data up to June 3, 2020, as can be viewed in Table 7, the predictions still got bettered. However the predictions are lesser accurate for USA where a spike in the cases reported near June 20, when compared to India.

**Table 5.** Predictions for the date June 20, 2020 with parameters based on data up to April 20, 2020.

| Country | Total cases |  | Active cases |  | Active+dead cases |  |
| --- | --- | --- | --- | --- | --- | --- |
|  | Actual | Predicted | Actual | Predicted | Actual | Predicted |
| Italy | 238275 | 209461 | 21212 | 30324 | 55822 | 69218 |
| India | 411727 | 75527 | 170269 | 26214 | 183546 | 33882 |
| USA | 2330578 | 1651967 | 1235657 | 794008 | 1357637 | 1077341 |
| Germany | 191216 | 159288 | 7555 | 3035 | 16516 | 4124 |
| Canada | 101019 | 57710 | 29121 | 5098 | 37531 | 7378 |

**Table 6.** Predictions for the date June 20, 2020 with parameters based on data up to May 20, 2020.

| Country | Total cases |  | Active cases |  | Active+dead cases |  |
| --- | --- | --- | --- | --- | --- | --- |
|  | Actual | Predicted | Actual | Predicted | Actual | Predicted |
| Italy | 238275 | 227266 | 21212 | 32481 | 55822 | 63546 |
| India | 411727 | 287805 | 170269 | 129296 | 183546 | 140072 |
| USA | 2330578 | 1868508 | 1235657 | 1013826 | 1357637 | 1153689 |
| Germany | 191216 | 171275 | 7555 | 1887 | 16516 | 3244 |
| Canada | 101019 | 98150 | 29121 | 17338 | 37531 | 24246 |

**Table 7.**
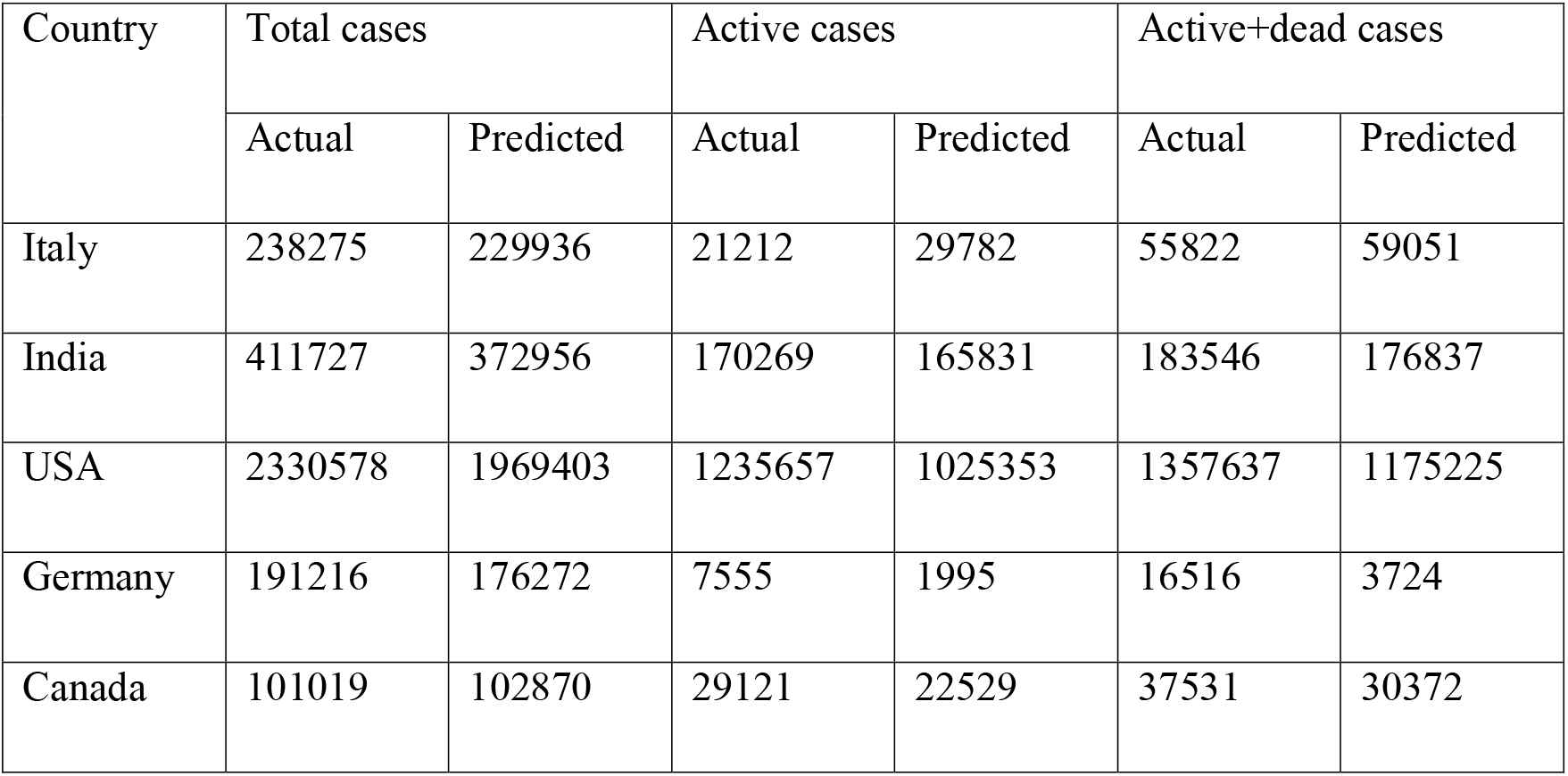
Predictions for the date June 20, 2020 with parameters based on data up to June 3, 2020.

